## Appendix 1 for "Organizational models and patient-reported outcomes for palliative care across five tertiary hospitals in Nigeria: an Environmental Scan"

| **S/N** | **Item** | **Yes** | **No** | **Ongoing** |
| --- | --- | --- | --- | --- |
| **1** | **Management buy-in/ approval and documentation** |  |  |  |
| 2 | **Infrastructure** |  |  |  |
|  | ***Service provision models*** |  |  |  |
|  | Home-based care |  |  |  |
|  | Inpatient/ outpatient hospice unit (space available for outpatient clinics and team/ |  |  |  |
|  | family meetings?) |  |  |  |
|  | Hospital Ward based services (ward rounds) |  |  |  |
|  | Operating days and hours decided? |  |  |  |
|  | **Building identified** |  |  |  |
|  | Is the building space adequate? |  |  |  |
|  | Are there rooms designated for the following (state number): |  |  |  |
|  | Waiting room |  |  |  |
|  | Consulting rooms |  |  |  |
|  | Pediatric waiting/playroom |  |  |  |
|  | Record room |  |  |  |
|  | At least one toilet facility |  |  |  |
|  | Administration Office |  |  |  |
|  | HOD office |  |  |  |
|  | Nurses’ room |  |  |  |
|  | Dressing/injection /medication dispensing room |  |  |  |
|  | Counseling room |  |  |  |
|  | Seminar room |  |  |  |
| 3 | **Personnel/ workforce strength** |  |  |  |
|  | **Is a Draft organogram available**? |  |  |  |
|  | State the number of personnel in each category | Number | How | How many |
|  | **Non-clinical**: |  | many | planned to |
|  | Health/ward assistants |  | trained | be trained |
|  | Cleaning staff |  |  | in 2022 |
|  | Security |  |  |  |
|  | Records officers |  |  |  |
|  | Unit secretary/ front desk officer |  |  |  |
|  | Accounts officer |  |  |  |
|  | Social Workers |  |  |  |
|  | Driver ( ) |  |  |  |
|  | **Clinical:** |  |  |  |
|  | Pharmacist ( ) |  |  |  |
|  | Nurses ( ) |  |  |  |
|  | Doctors ( ) |  |  |  |
|  | Physiotherapists ( ) |  |  |  |
|  | Volunteers **( )** |  |  |  |
| 4 | **Training/ certification**  Draft schedule for weekly team seminars available?  Any certificate training/workshop plan for personnel in 2022?  Any plan for conference attendance by members of staff? |  |  |  |
| 5 | **Communication** |  |  |  |
|  | Dedicated Unit phones |  |  |  |
|  | Printers |  |  |  |
|  | Stationery |  |  |  |
|  | Scanners/ photocopiers |  |  |  |
|  | Cabinets |  |  |  |
|  | Games |  |  |  |
|  | Chairs, mats |  |  |  |
|  | TV |  |  |  |
|  | Laptops |  |  |  |
|  | Internet connection |  |  |  |
|  | IEC materials for clients containing unit contact details, clinic times, and services |  |  |  |
|  | provided |  |  |  |
| 6 | **Patient Records Management** |  |  |  |

|  | Case files- with printed care questions covering e.g., social, spiritual topics, plan of care, bereavement plans  Enrollment cards  Tablets for electronic record management Appointment system scheduler  Referral sheets |
| --- | --- |
| 7 | **Transportation**  Bus/SUV for rounds and home visits  Links to an ambulance service |
| 8 | **Finances** Fuel Stationeries  Miscellaneous  Receipts and invoices for client visits: home and outpatient  Dedicated unit bank details for clients |
| 9 | **Image/awareness creation**  Signages at the unit e.g., signboard  Information fliers to be distributed in the hospital A memo to heads of department  Plan for a grand round to notify health care providers in the hospital |
| 10 | **Data management and research**  Data entry dashboard for all patients seen e.g., excel Research and publication plans for the unit are available |
