## Appendix 2 for "Organizational models and patient-reported outcomes for palliative care across five tertiary hospitals in Nigeria: an Environmental Scan"

| **Survey Population** | | | |
| --- | --- | --- | --- |
| Leadership  (Physician Leads, Administrators) | Frontline Workers (Nurses, Physicians, Social Workers) | Adult Patients | Caregivers |
| **Survey Content** | | | |
| - Infrastructure - Resources - Operations - Staffing - Training - Patient profiles | - Training - Expectations and Attitudes toward PC - Desired Knowledge | - Pain - Symptoms - Worry - Sharing feelings - Will to live - Peace - Planning for future | - Information regarding the received care - Confidence in caring for patient - Worry about the patient |
| **Assessed with** | | | |
| Multiple choice, checkbox, and free response question survey | | 5-point Likert scale survey | |
| **Informed by** | | | |
| Consolidated Framework for Implementation Research (CFIR); Centre for Palliative Care in Nigeria (CPCN) Checklist | | African Palliative Care Association's African Palliative Outcome Scale | |

**Environmental Scan Survey Summaries**
